# Tackling the first COVID-19 wave at the Cape Town Hospital of Hope: Why was it such a positive experience for staff?

**DOI:** 10.1101/2022.02.21.22271306

**Authors:** Steve Reid, Mitan Nana, Theo Abrahams, Nadia Hussey, Ronit Okun-Netter, Tasleem Ras, Klaus von Pressentin

**Affiliations:** Primary Health Care Directorate, University of Cape Town; Dean’s office, Faculty of Health Sciences, University of Cape Town; Western Cape Department of Health; Division of Family Medicine, University of Cape Town

**Keywords:** COVID-19, intermediate care facility, field hospital, psychological resilience, professional burnout, health team, interdisciplinary, health care team, culture, organizational, leadership

## Abstract

**Background:** In contrast to alarming reports of exhaustion and burnout amongst healthcare workers in the first wave of the COVID-19 pandemic, we noticed surprisingly positive staff experiences of working in a COVID-19 field hospital in South Africa. The 862-bed ‘Hospital of Hope’ was established at the Cape Town International Convention Centre specifically to cope with the effects of the first wave of the COVID-19 pandemic in Cape Town.

**Methods:** We aimed to systematically describe and assess the effects on staff and the local health system. A cross-sectional descriptive study design was employed using mixed methods including record reviews and interviews with key informants.

**Results:** Quantitative results confirmed high job satisfaction and low staff infection rates. The emerging themes from the qualitative data are grouped around a “bull’s eye” of the common purpose of person-centredness, from both patient and staff perspectives, and include staff safety and support, rapid communication, continuous learning and adaptability, underpinned by excellent teamwork. The explanations for the positive feedback included good disaster planning, adequate resources, and an extraordinary responsiveness to the need.

**Conclusions:** The ‘Hospital of Hope’ staff experience produced significant learnings for the design and management of routine health services outside of a disaster situation. The adaptability and responsiveness of the facility and its staff was largely a product of the unprecedented nature of the pandemic, but such approaches could benefit routine health services enormously, as individual hospitals and health facilities realize their place in a system that is ‘more than the sum of its parts’.

## INTRODUCTION AND BACKGROUND

An 862-bed COVID-19 field hospital, the ‘Hospital of Hope’ (HoH), was established at the Cape Town International Convention Centre (CTICC) as an intermediate care facility specifically to cope with the effects of the coronavirus disease 2019 (COVID-19) pandemic in Cape Town, South Africa. The temporary HoH was commissioned by the Western Cape Provincial Government on 1 April 2020 to meet the anticipated pressure on the acute hospital services in the city during the imminent first wave of the COVID-19 pandemic. The overall aim of the hospital was “to offer hope by delivering high quality, efficient inpatient care in response to the needs in the Cape Town metropole, while ensuring the safety and positive growth of staff.”

By 14 August 2020 medical care had been provided to 1502 patients, referred from other healthcare facilities in and around Cape Town. The 862-bed HoH facility accepted patient admissions for only a 10-week period (8 June 2021 – 14 August 2021), which saw 303 transfers out of the facility to higher levels of care and 81 in-patient deaths. The inpatient beds were organised into three main patient categories, namely those patients diagnosed with COVID-19 requiring inpatient oxygen therapy, those patients diagnosed with COVID-19 which complicated their underlying chronic conditions, and those patients with COVID-19 deemed for palliative care based on the PALCARE guidelines.^1^

Much of the evidence currently available reports the experiences of staff working at established hospitals, whereas for field hospitals reports are lacking. An Australian study^2^ found that the pandemic had a significant effect on the psychological well-being of hospital clinical staff: most were concerned about contracting COVID-19, infecting family members and caring for patients with COVID-19, but positive aspects were also described. However, the literature has so far offered limited insight into the experiences of staff stationed at dedicated field hospitals during the COVID-19 pandemic. A qualitative study at Nightingale Hospital in Manchester reported that doctors had an overwhelmingly positive experience.^3^. There was consistent mention of a strong team; in particular the feeling of being individually valued within a flattened hierarchy. Staff wellbeing and education were also regularly mentioned and helped contribute to this overall feeling. When asked what they would take forward, doctors focussed on the importance of a strong team that values multidisciplinary working. But the hospital was not without challenges, with processes changing from one shift to the next and leading to potential errors. In addition, system issues such as with medication and documentation led to a sometimes chaotic work environment. Staff identification was a significant challenge, and potentially contributed to communication breakdowns.

In view of the anecdotal feedback from HoH staff indicating that the experience of working at this field hospital was extraordinarily positive, we aimed to describe and assess the staff experiences and effects on the health system of the temporary HoH during the height of the first wave of the COVID-19 pandemic in Cape Town, from the perspective of staff members and health managers.

## METHODS

### Study design

A cross-sectional descriptive study design was employed using mixed methods including record reviews of existing documents, and interviews with key informants.

### Study setting

The population of the Cape Town metropole is served by both public and private sector health services. The majority of the 4 602 248 population have access to basic municipal services (service coverage of 96.4% for water, 94% for refuse removal, 93.7% for electricity, 90.9% for sanitation and 77.2% for housing). The public sector consists of 126 primary health care facilities, which drain to several district, regional and tertiary hospitals. These acute hospitals at regional and tertiary levels have high care and intensive care facilities available, including ventilators and specialist staffing. The two tertiary hospitals (Tygerberg and Groote Schuur hospitals) drain the whole Western Cape provincial public sector service.

### Study population

The HoH staff members consisted of 463 clinical staff (52 medical officers, nine medical specialists, 360 nurses led by eight operational managers, eleven physiotherapists, four dieticians, eight pharmacists, ten post basic pharmacists, four radiographers and three social workers) and 210 non-clinical staff (administrative, supply chain, porters, housekeeping and catering staff), as well as an operational management team. During the commissioning of the HoH, 722 posts were approved and almost 400 of these were filled in record time. At first there were insufficient nursing operational managers and certain administrative staff such as admissions clerks, an infection control coordinator, and an occupational health coordinator. These posts were filled during the remainder of the operational phase to give a total of 673 personnel.

Qualitative and quantitative data was derived from four sources of existing records, which were reviewed to answer the research question:

a. A voluntary and anonymous exit survey was administered to all staff on leaving the facility as it was decommissioned including quantitative and qualitative responses. The staff exit survey was developed by the researchers, made available online and all staff members were encouraged to complete it (*Appendix 1*).
b. Electronic records from the Vula Mobile referral app^4^ were used for care coordination and transfers between the acute hospitals and the HoH.
c. Minutes of all management meetings held during the operational phase of the HoH, including a close out meeting held at the end of the 10-week period during which managers and clinicians from the acute hospitals gave feedback to the HoH operational management team.
d. The COVID-19 infection occupational health records of all staff.

Key informant interviews were held with purposively selected individuals including HoH team leaders, the HoH management team, managers of other health facilities in the Cape Town metropole, as well as head office managers who planned the HoH, using a semi-structured interview guide developed by the research team (*Appendix 2*).

In terms of data collection, the records were readily available to the research team. For the key informant interviews, a maximum of 12 potential participants were purposively selected and invited to be interviewed by virtue of their involvement in the HoH, of which all 12 participated. These interviews were held by telephone, digital communication (such as Zoom^5^) and in-person, recorded and professionally transcribed verbatim. The records and interview transcriptions were collated and safeguarded by the research team.

With regard to data analysis, quantitative data from the records were entered into Microsoft Excel Spreadsheets^6^ and simple descriptive statistics were used to analyse the data and to generate tables and figures. Responses to open-ended questions in the exit survey as well as the minutes of management meetings and key informant interviews, were analysed by thematic analysis using the framework method^7^ to produce major and minor themes in relation to the research question.

In terms of reflexivity, the research team consisted of members of the HoH operational management team (SR, TA, RO, TR, KvP), a HoH clinician (NH) and a research officer employed at the UCT Faculty of Health Sciences deanery (MN). SR, TR and KvP are academic family medicine specialists employed by the University of Cape Town, RO is a family medicine specialist, TA is a physical rehabilitation therapist and health service manager, and NH and MN are both early career medical doctors.

### Ethical considerations

Exit surveys were anonymous at the level of data entry. Data from Vula Mobile App,^4^ staff leave records and management meeting minutes were rendered anonymous during the process of data collation and analysis. Confidentiality of participants in interviews was maintained by excluding names and identifying details from the transcripts and substituting participant codes. The protocol was approved the Human Research Ethics Committee at the University of Cape Town (reference 503/2020), and permission to conduct the study was obtained from the Western Cape Department of Health through the National Health Research Database (reference WC_202010_037).

## RESULTS

The staff exit survey was completed by 190 staff members giving a response rate of around 30%. There was a good response rate from all professional staff except for nursing staff. Two-thirds of the medical officers and Cuban doctors completed the survey. 100% of consultants (emergency, internal medicine, family medicine) completed the survey. The survey was completed during the last week before the hospital was closed, by which stage many nurses had been relocated to other work, which may account for the low response rate from the nursing staff. The length of experience amongst the respondents ranged from less than 1 year to over 15 years. A quarter of the respondents were not from the Western Cape. The results however reflected there were no significant differences in responses given irrespective of place of origin.

In addition to the qualitative data from the exit survey, 12 key informants were interviewed.

The findings and main themes from the mixed methods sources may be organised according to the key aspects of the logic model, as illustrated in Figure 1.

**Figure 1:**
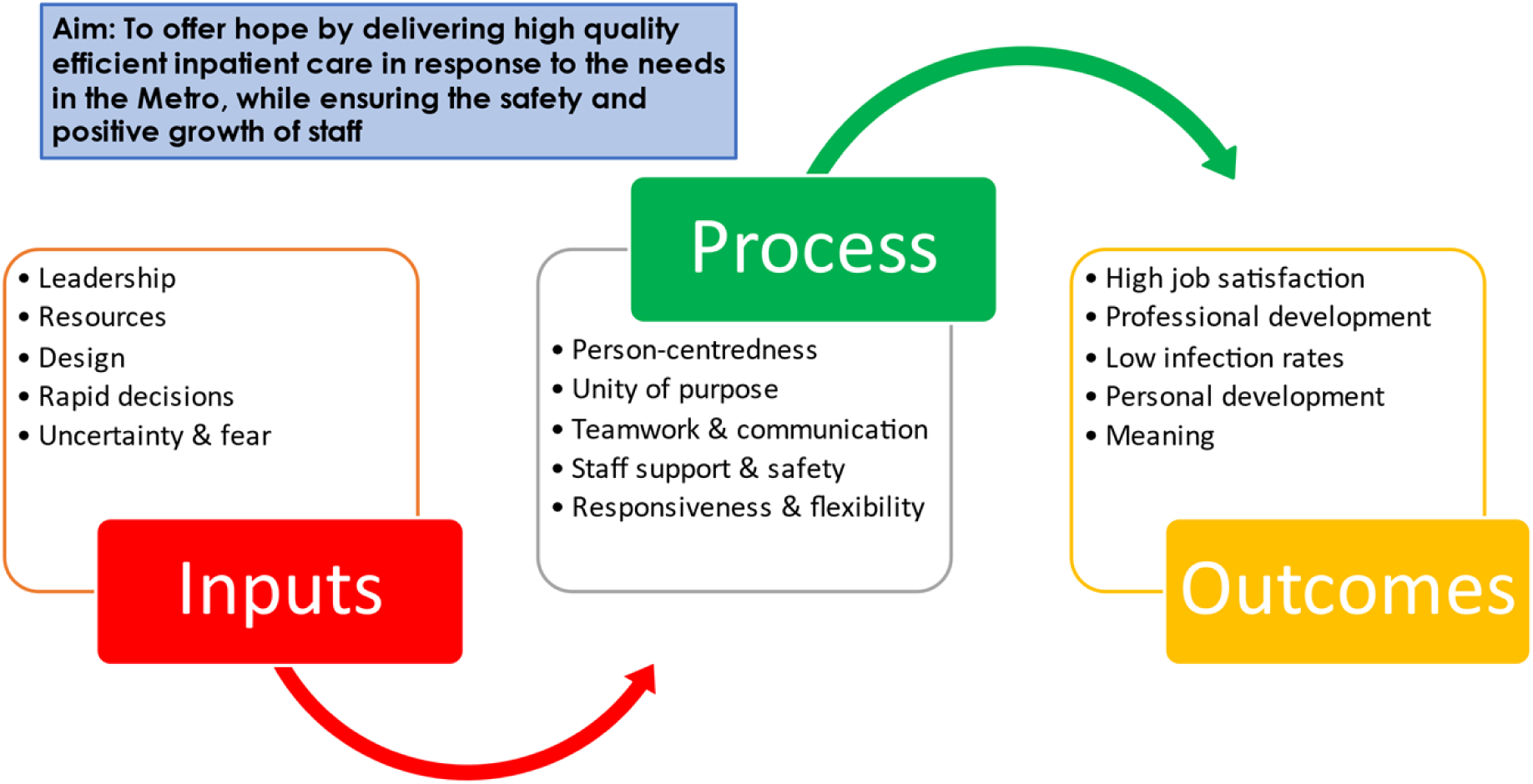
Overview of themes

### 1. Inputs: Design and set-up

It was clear from the interviews with managers, that a significant level of leadership was demonstrated in the early phases of the pandemic in March 2020, when high-stakes decisions had to be made quickly, and significant resources committed in an extremely uncertain context. Putting together a dependable management team in a hurry was initially daunting for the senior manager involved, but he relied on previous experience:

> *“To be honest the first impression I had was this is not a do-able task and when you are faced with an undoable task you turn to people that have done undo-able tasks for you previously*.*”* (Senior manager)

Pre-existing relationships of trust between managers played a major part in this process, as one put it:

> *“I must say clearly that the reason we could do that was because we trusted that the system would have our back if something went wrong*.*”* (Senior manager)

The responses of managers and senior clinicians was encouragingly positive, and this spirit of wanting to contribute despite the fear and the risks involved, was echoed by the staff who were recruited.

> *“I think when senior people put up their hands it kind of makes it easier to attract a team around them*.*”* (Manager)
>
> *“The opportunity to work with ambitious healthcare professionals was amazing”* (Manager)
>
> *“We were all facing new thing nobody have experience in COVID, from professional nurse to nursing assistant, we were all the same. What a wonderful moment”* (Senior nurse)
>
> *“Especially getting a chance to work with the Cuban doctors. Even though there was a language barrier … it was a privilege to work together and demonstrate international solidarity to battle the pandemic”* (Senior clinician)
>
> *“Being part of history and contributing to the country’s time of need”* (Manager)
>
> *“Unity. All working together to reach one goal”* (Senior clinician)

In terms of resources, estimating upfront the number of beds that would be required at the peak was a tricky decision that was led by epidemiological models predicting enormous numbers of patients. In retrospect, opening 862 beds with 8 high-flow beds in CTICC1 proved to be the right decision, as a maximum of 325 normal beds and only three of the high-flow beds were occupied at any one time at the peak. As the numbers of patients increased, there were plans and pressure to expand the facility into CTICC2 by a further 400 beds, but again the right call was made under pressure, demonstrating courageous leadership:

> *“There was also a narrative in the department to open CTICC2 …. because that was agreed to by the department - ready to roll kind of thing and …… I took it back to Texco* [Western Cape Provincial Department of Health’s Top Executive Committee] *I think a total of three times to say you guys are making a mistake, this is not the right thing”*. (Senior manager)

The deliberate design of the hospital infrastructure to prioritize infection control by controlling staff and patient flow, played an important role in reducing cross-infections and creating efficiencies, in addition to the high roof and air-conditioning of the conference halls which aided ventilation. The design of the facility appropriately involved infectious disease and emergency medicine consultants with field hospital experience, who worked at great speed to create systems and flows from scratch. When the first patient arrived however, the medical management was deliberately handed over to family physicians, as appropriate to the intermediate level of care. This created a discontinuity in the context of the high pressure of commissioning the hospital in time, which resulted in some conflict:

> *And so I think that was very challenging for the new people coming on and it was also challenging for us who had been there and had this momentum and wanted to go fast and everyone else needed time to adjust. So there was quite a bit of conflict as you would imagine with something of this scale with so many people from different backgrounds involved*. (Senior clinician)

The medical management of the facility largely by family physicians however, proved to be justified in retrospect, as a person-centred approach to both staff and patients seemed to make a significant difference to their respective experiences (see *Outcomes* below).

> *I didn’t expect such a primary care family medicine leading orientation, but I think it is working incredibly well there is no criticism at all but the way the care was managed and the family medicine approach I think is very different to the internal medicine approach*. (Senior clinician)

### 2. Process (Implementation)

#### a. Person-centredness

Many staff members were surprised by the experience of holistic patient care using the family medicine approach and realized its value in retrospect.

> *“Having a largely “Emergency Medicine” background, one often neglects holistic care and focus on a problem orientated approach to clinical medicine. Working at the CTICC has been an interesting experience in this regard as on more than one occasion, we have seen significant focus on non-clinical components to health care which has invariably and often somewhat surprisingly led to some incredible recoveries. It has made me reconsider and re-value the importance of holistic medicine/care and many other family medicine principles which one often takes for granted in a busy clinical setting”* (Senior clinician)

Staff developed closer relationships with patients than they had expected, due to the nature of the care required, particularly in palliative care. This also influenced the experience of teamwork, as articulated by the following respondents:

> *“I didn’t expect such a family experience, all disciplines came together to learn from one another. Mostly the hierarchy fell away*.*”* (Clinician)

#### b. Unity of purpose

At the onset of the pandemic when HoH was still being commissioned, there was a great deal of fear: fear of the unknown; “fear of missing out” on being on the frontline, combating COVID-19 and preventing illness and death; and fear of contracting and succumbing to COVID-19 itself. Although the level of fear gradually dissipated as the pandemic wore on and more knowledge emerged, it must be acknowledged that *“people were actually very fearful. There were degrees of it and people expressed it in different ways*.*”* (Manager)

However, stronger than that fear was the *unity of purpose* – a purpose to “contribute to humanity and the country as a whole” and “do the right thing”; a purpose to answer a call during a time of “great need”; a purpose to learn and grow both personally and professionally; and a purpose to be involved in something new, exciting and possibly “an opportunity of a lifetime”.

This unity of purpose inspired compassion, extreme diligence and hard work, great personal sacrifice. A culture of comradery and a ‘can-do’ attitude was born even among staff who were unfamiliar with each other and where in some cases had no previous experience.

> *“When it comes to saving lives and helping people… people will go that extra mile regardless of who they are or where they come from…”* (Senior clinician)
>
> *“People from all the different angles… [those with] clinical experience, [those at a] managerial level, people that have got only administrative backgrounds, people with certain expertise… just going beyond what fits in their original scope… There was this common goal and everyone put in a hundred and fifty to two hundred percent… Everyone that was involved in the project was just ready to deliver … putting the best possible COVID response in place and that was so refreshing*.*”* (Manager)

There was an overwhelming sense of “togetherness”. It seemed that *“at the end of the day everybody was just trying to do their best and everybody was passionate about what they were doing and trying to get the right thing done for their patients*.*”* (Senior clinician)

One respondent even said that: *“Leaving home coming to work made me feel like a hero knowing I’m coming to assist in this difficult time that the country is facing. Helping a patient when ever [sic] I was doing my rounds in the wards came easy to me it really woke up the spirit of Ubuntu*.*”* (Clinician)

#### c. Teamwork and communication

The team-dynamics were informed by a culture supportive of teamwork, as well as the necessary resources to encourage this style of organisational culture. In terms of leadership, the respondents noted the approachability and transparency of the management teams: *“The doors were always open to speak about how we felt at any given time”* (Senior clinician). There was also a level of honesty and humility displayed by managers. Managers were *“honest about the fact they [they] did not have all the answers”* (Manager) but there was a call to collaborate and work together. In turn, this display of honesty and humility may have *“put people at ease and they were more willing to come up with innovative ideas and [to] contribute”* (Manager). While no explanations could be offered, a sense of trust was quickly established among the different managers despite many not having worked together previously and the high stress levels associated with the pandemic and setting up a facility from scratch in such a short space of time. This trust appeared to have increased the level of collaboration and led to lower levels of disappointment and higher levels of forgiveness when expectations were not met, or mistakes were made.

As far as the clinical staff were concerned, a flattening of the hierarchy and the provision of equal opportunities to participate and contribute within teams led to team members feeling valued, respected and validated as team members: *“there was true interdisciplinary teamwork”* (Senior clinician). This was echoed by a survey respondent: *“The environment was very conducive to learning, not hierarchical, completely patient centred and everyone was friendly*.*”*

In the survey, many responses emphasised the collaborative nature of inter-disciplinary teamwork and connection, and the term “family” was used often. *“I didn’t expect such a family experience, all disciplines came together to learn from one another. Mostly the hierarchy fell away*.*”* Other respondents noted equally positive responses, including teamwork, approachable consultants, the positive attitude of all staff, interdisciplinary cohesion, and high emotional support for staff (e.g. wellness area, debriefing). One young pharmacist described experiencing more integration in the team and clinical context, *“As a newly registered pharmacist, …. the Hospital of Hope allowed me to experience an interdisciplinary team like never before. Being able to work directly with patients gave me a new sense of passion for my profession by allowing me to see the direct result of my influence*.*”*

In addition to the transparency, honest and humility displayed by management, communication was regular, open and timeous: “*every single day there was constant communication with every team*” (Manager). Respondents commented that this form of communication led to the creation of “*safe [working] spaces*” in which issues and feelings could be communicated, particularly during a time of intense fear, uncertainty and stress. The effective communication even extended to “*between hospitals [that helped to create a feeling that we are all in this together*” (Senior clinician, close out meeting). This important means of communication were the “*daily huddles*” via Microsoft Teams^8^ with management representatives of all the acute hospitals in the Cape Town metropole. These huddles were described as “*short, to the point, informative… [everyone was] on the same page at the same time in the day… it was [an] extremely effective [means of communication]”* (Senior manager at referral hospital, close out meeting).

One of the key informants noted, that “*there were people who left - there were staff who left they were unhappy, they felt like they were working too hard and that the place was not running well*.” (Senior clinician). The survey respondents also described some challenges relating to teamwork and communication. Some of these challenges may relate to the perceived fluid state of organisation, especially during the early operational period: “*Earlier designation of team leaders*”; “*Better organisation of processes on the ward such as for diabetics, dispensing of meds by nurse*”; “*Better role clarification*”; “*Even though I’m a cleaner I also did other roles such as hostess. It would be better if there was more clarity as to what my role is from the very beginning*”; “*There could’ve been more support on the floor with operational issues. Like where to find stock, office allocations and such*”; “*Multidisciplinary strategic planning could have been filtered down to subordinates better*”; “*Initial communication channels took long to be formatted-could have been set up from get go - but once set up was easier to channel issues*.”

#### d. Staff safety and support

A remarkably low level of staff infections with COVID-19 was found. As shown in Table 1, 2.4% of staff tested positive for COVID-19 over a two-month period, compared to an infection rate of 8.1% amongst staff over the same period at a nearby hospital of equivalent size.

**Table 1:** Staff infections with COVID-19

| Variable | CTICC HoH | Comparable hospital |
| --- | --- | --- |
| Total number of staff on 31 July 2020 | 673 | 795 |
| Total staff COVID-19 positive between 8 June and 14 August 2020 | 16 | 64 |
| Health Professionals who tested COVID-19 positive | 16 | 41 |
| Non-professional staff who tested COVID-19 positive | 0 | 23 |
| Total staff positivity rate | 2.4% | 8.1% |

Despite initial concerns over personal protective equipment (PPE) and isolated incidents during which PPE was unavailable, overall staff reported feeling safe. Several reasons were offered. First, knowing that all patients were COVID-19 positive acted as a constant reminder of the risks of transmission, and maintained a heightened sense of awareness and caution. Second, the high ceilings of the Convention Centre contributed to good ventilation. Except for isolated cases, there was generally adequate PPE; the proper usage of which was monitored by dedicated personnel. Third, posters positioned at key points within the hospital served as a constant reminder to use PPE correctly and appropriately, and appropriate occupational health and safety training was performed as well as systems and protocols implemented through daily team huddles, and staff were encouraged to eat well and consume vitamin supplements to boost their immune systems. Stationery and consumables could not be moved from one ward or section of the hospital to another, and mobile phones and devices had to be kept and used within clear plastic bags to prevent fomite transmission. Following the first incident during which there was a shortage of scrubs, various contingency measures were implemented including encouraging staff to carry two sets of clothing and the use of jump suits.

The survey results reflect a uniformity in perceptions of a high level of support provided at HoH amongst all cadres of staff, as reflected in Figure 3. More than 80% of staff felt safe at work, and the majority felt they could access assistance with emotional support when they needed it. Over 80% felt appreciated and valued. The majority also responded that they were able to access assistance with solving problems when the need arose. Only the responses about the debriefing sessions were found to vary amongst the staff with pharmacists finding the debriefing sessions to be less helpful than the other cadres of staff. The debriefing sessions were also found to be more helpful amongst the more experienced staff (> 11 years work) than the less experienced staff (<1 year).

In qualitative feedback, respondents reflected that morale was high despite the experience of establishing HoH as a *“time of very hard work… trying to put something in place in such a short period, something that didn’t exist and then that was then established in such a short time”* (Manager). Also, staff absenteeism was at an absolute minimum with *“very few staff staying away from work unless they were really, really sick…*” (Manager). Complaints about staff, although present, were also scant. Moreover, managers reported that teams did not *“require micro-management… [One could] just stand back and let them do the work*” (Manager).

It also appeared that staff felt supported, not just physically and with their work but emotionally too. Working at Hospital of Hope was *“quite an emotional rollercoaster”* with high levels of stress for some but overall, it appeared that staff felt “protected” and supported and that this may have further contributed to “just giving [their] all”. It was reported that key members of management were often seen on the floor supporting the staff and expressing concern over their needs and emotions with ‘check-ins’ occurring daily in most cases. One manager remarked that *“because [he was] showing [his] face down there, [he was hopeful] that that also gave them the assurance that [the staff] are not left alone that [he was] there with them supporting them as well”* (Manager)

> *“Being surrounded or working with a group of people who were so committed and also determined to make a difference, the healthcare team, the admin team, the management, the volunteers, having debriefing sessions and I do think that the platform was there for myself and my team to feel safe and feel supportive and also we weren’t forced to do anything that we didn’t want to do*.*”* (Clinician)

Staff support initiatives seemed to have also contributed to the feeling of being supported and to the high levels of morale. These included the various staff wellness initiatives such as having a senior person on the clinical management team dedicated to staff wellness who was available to staff to discuss any issues they may have had and who co-ordinated the various staff wellness initiatives such as: debriefing sessions; community volunteers distributing gift packs to staff; the ‘Handwashing’ song (see https://youtu.be/bQCOVNdoYjM); a staff ‘Wellness hub’; and a staff appreciation ceremony. Other wellness initiatives included singing and recording the ‘Jerusalema’ dance challenge as a team (see https://www.facebook.com/watch/?v=228267391738155), a common sight during the pandemic at many South African hospitals; and “the other extras. *“It really made the place feel like a family!”* (Clinician)

In the survey, staff members felt more protected by the PPE than they expected as evidenced by one staff member saying that: *“I was expecting to see some of us contracting the virus, but the government saw to it that we are protected*.*”* Another staff member said that: *“I thought I would be affected too but because of the proper PPE I have no fear and I thank you for that”*. In the survey, it appears that their fears and expectation of danger related to working with COVID-19 patients changed during their experience at HoH. One staff member commented that *“At first before coming to work at CTICC I was scared to come because I thought I’m standing a chance of getting infected by the COVID-19 but as time goes on, I felt safe and free at work and I just became very grateful to be working at CTICC*.*”*

Some survey respondents offered critique: *“There was no support at all we were ignored by management because we’re working nights shift, the don’t care about night staff”; There could’ve been more support on the floor with operational issues. Like where to find stock, office allocations and such*.*”*

#### e. Responsiveness and flexibility

Despite the many challenges faced, including the rapid evolution and unfolding of events, a lack of clear reporting lines, roles and responsibilities, and people with differing levels of experience, a novel degree of flexibility and adaptability was displayed in the way Hospital of Hope was able to respond and adapt to the needs of the broader health platform during the first wave of the pandemic. Not only was the Hospital of Hope able to *“relieve pressure from the system… and support the system”* (Manager), but there was also a consensus by other institutions that its opening and role in the system was *“like the cavalry coming over the hill to save them*”. (Manager). A senior clinician at the close out meeting noted that, *“[it was the] responsiveness to the need that stood out for me*.*”*

While Hospital of Hope added extra beds to the platform as a whole, which *“unloaded some of this high pressure”* and helped clear overflowing wards and corridors at some of the other facilities, the *“positive effect that it had on the service platform [was] because of its responsiveness”* (Senior manager) and the fact it *“had a platform view”* (Senior clinician).” Management of other institutions on the platform remarked that they had felt supported by the Hospital of Hope in the way it was able to accommodate their changing needs in real time through effective, daily communication across the platform. Rarely were patient referrals rejected and a feeling that “we were all in this together (Senior clinician, close out meeting)” permeated through the system. Hospital of Hope had such a meaningful impact that its closing was described as the end of “*our period of respite* (Senior clinician, close out meeting)” despite being amidst the pandemic.

Moreover, it was noted that Hospital of Hope “*challenged the dogma of how we provide health care… [There was an] ability to share resources across the platform based on need and access to healthcare [Clinical manager, close out meeting]*”. This led to a greater “*sharing of resources between primary health care, district level hospitals [and] tertiary level hospitals* (Clinical manager, close out meeting)”.

There appeared to be an openness and willingness, on the part of individuals and the institutions, to deviate from the traditional, sometimes deeply established “*ways of doing things*”. In the words of one respondent, “*things were happening very quickly and in a way that you never saw happen in the Department of Health… There was this great willingness to get things done and then just an openness*… (Senior clinician, close out meeting)”. People had somewhat accepted that they work “*for a bigger department and not on a small island… [There was an openness] to the idea that we can support the platform and not just ourselves* (Manager).”

Of course, it is also possible that the flexibility and responsiveness was due to the rapid pace of events. *“Everything just happened so fast… there was almost no time to think about it too much. I think we were all on this rollercoaster ride*.” (Manager). One respondent remarked that *“We could have really played hard ball and just said listen here, we are not up for this and these are the boundaries and we are reinforcing these boundaries* [Manager]”. However, a very different approach had been adopted, which was echoed during the closeout meeting: *“Responsiveness… There was some criteria for admission… [It was] very quickly established that the criteria was not going to meet the needs [of the rest of the platform]… [There was a] rapid adjustment based on feedback [to meet the needs]*.*”*

### 3. Outcomes: “Hitting the target bullseye”

The key themes related to the implementation and factors that contributed to person-centredness were described by one key informant as ‘hitting the target bull’s eye’ (figure 2), a summary which served to frame the major themes arising from the data. At its core is the person-centredness approach which infused the daily activities linked to patient care and staff wellness.

**Figure 2:**
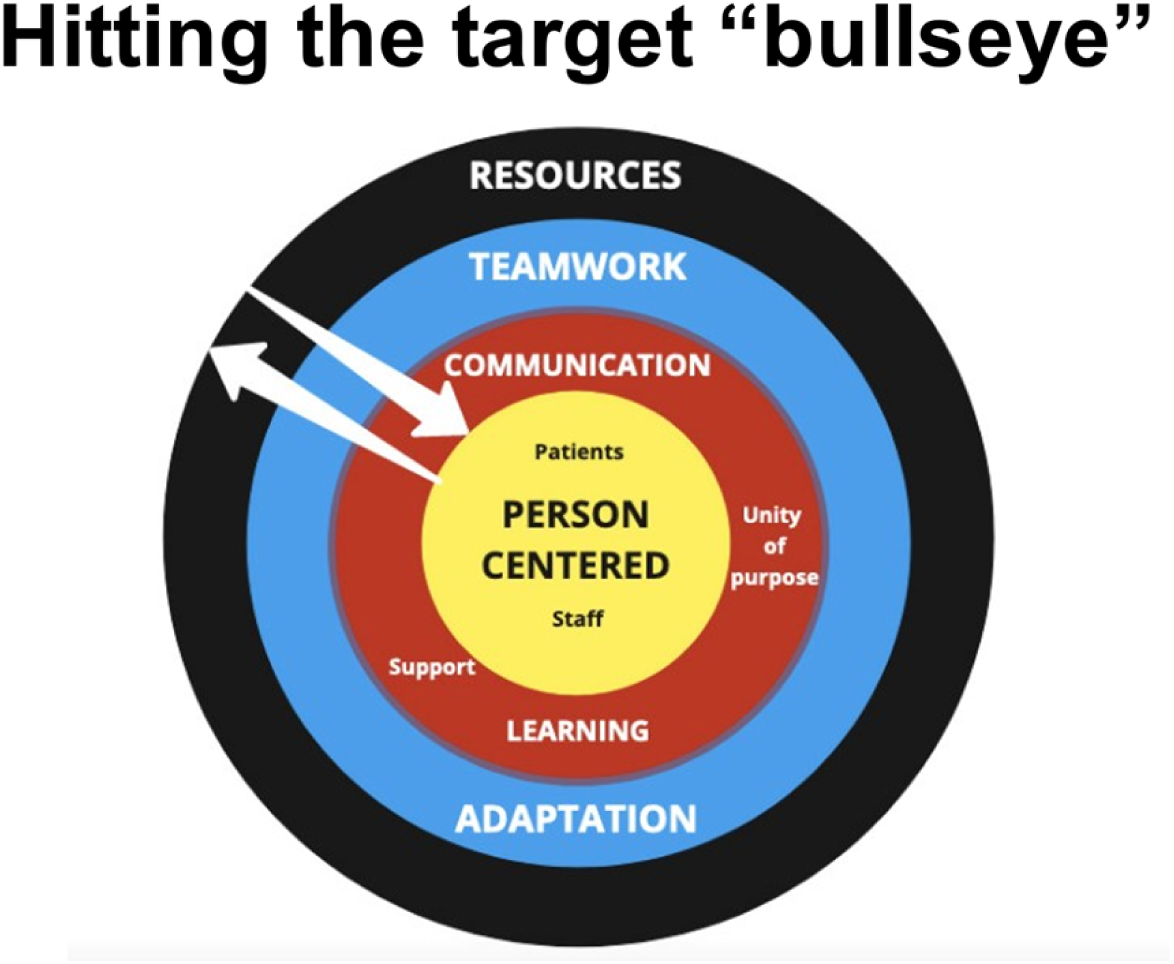
A Diagrammatic representation of the central process themes

> *“So reflecting on that, I think we really hit the target bullseye when we were able to deliver on those patient experiences and giving a good patient experience and I think some of the best comments that came through is where people started talking about hope and just their interaction with the staff and being able to be accommodated in that space and where it was really in the peak of the pandemic. They experienced it as a time of hope and not a time of despair and I think that was remarkable*.*”* (Manager)

A number of factors contributed to this sense of achievement, including high job satisfaction, professional and personal development, and discovering a deeper sense of meaning in the work. Finally, a significant outcome was the learning arising from this experience that can be taken into routine health services once the pandemic is over.

#### a. High job satisfaction

Staff were surprised by the positive nature of their experience, how much they enjoyed working there and how sad they were when it was over. There was an expectation that it would be busier and more stressful than it actually was. Staff developed closer relationship with patients and their families than they had expected. And many responses emphasised the nature of inter-disciplinary teamwork and connection, including words such as “*family*” often.

> *“There was a lot going right but unfortunately you don’t see what is going right, you just concentrate on that was going wrong because that is what is in your head - that is what everyone is complaining about, that is why people are screaming at you to fix. So all in all, it was a wonderful experience - completely life changing for me*.*”* (Manager)
>
> *“Despite all the stresses and challenges relating to the COVID response… During this COVID time, I’ve been more creative and productive than I was before then*… *there are aspects in my job that I… forgot about that I’m rediscovering and enjoying a lot”* (Manager)

Staff also felt more protected by the PPE than they expected they would be.

> *“I was expecting to see some of us contracting the virus, but the government saw to it that we are protected*.*”* (Nurse) *“I thought I would be affected too but because of the proper PPE I have no fear and I thank you for* that” (Clinician)
>
> Their fears and expectation of danger related to working with COVID-19 patients changed during their experience at HoH:
>
> “*At first before coming to work at CTICC I was scared to come because I thought I’m standing a chance of getting infected by the COVID-19 but as time goes on I felt safe and free at work and I just became very grateful to be working at CTICC*.” (Nurse)

#### b. Professional development

Generally, staff felt that the HOH experience would assist their future careers, and that their competence improved, as shown by the exit survey results reflected in Figure 3.

**Figure 3:**
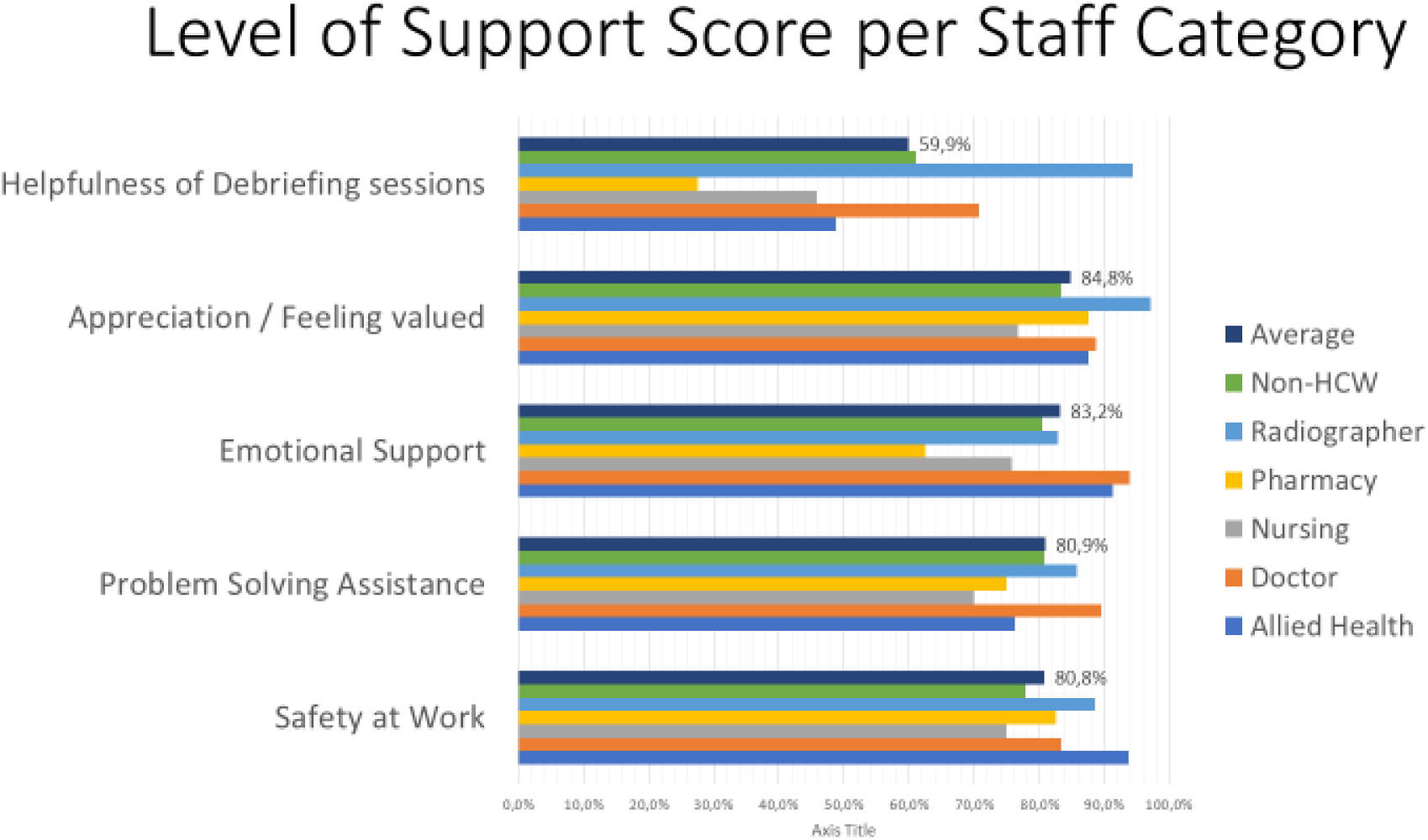
Level of Support Score per Staff category

An average of over 90% of staff felt more competent to do their work after working at HoH, and an overwhelming majority found working at HoH to be beneficial to their future careers. Areas found to be where most learning was achieved in descending order was from daily work (84%), discussions with peers (64%), in service training (47%) and then meetings or huddles (46%).

When asked to explain briefly to what extent this experience would assist their future careers, they spoke about leadership skills, self-confidence, experience in teamwork, the value of the multi-disciplinary team, better communication skills and networking, as well as clinical competence.

**Figure 3:**
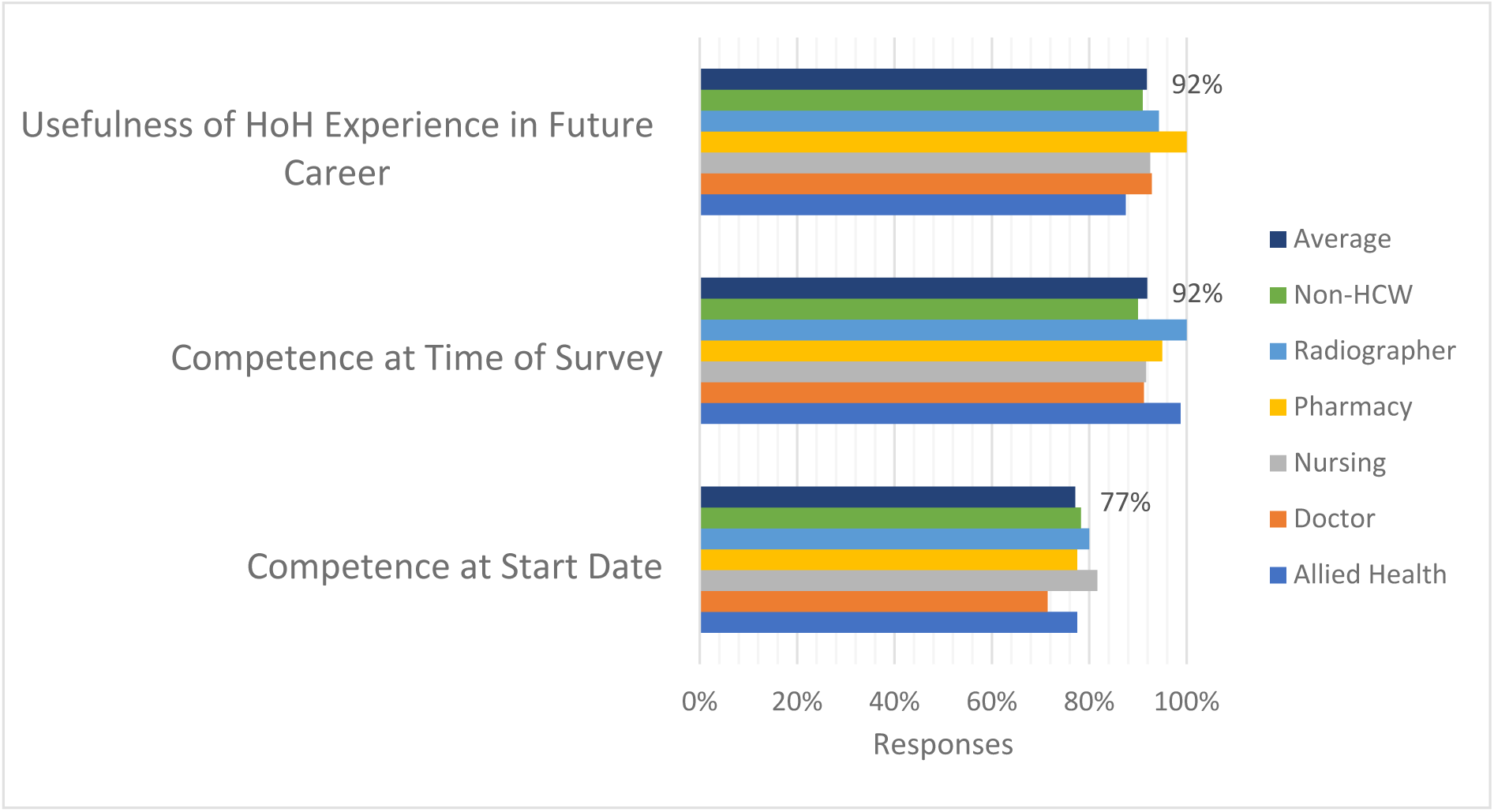
Reported professional development of staff

#### c. Personal development

Most staff enjoyed their work at HoH: on average across categories of worker, 92% of survey respondents (see Figure 4). More than 90% felt they grew personally through their experience at HoH, and less than 40% found going to work stressful. Over 90% found working at HoH meaningful and worthwhile, and 90% of staff reported that their teams worked well together.

**Figure 4:**
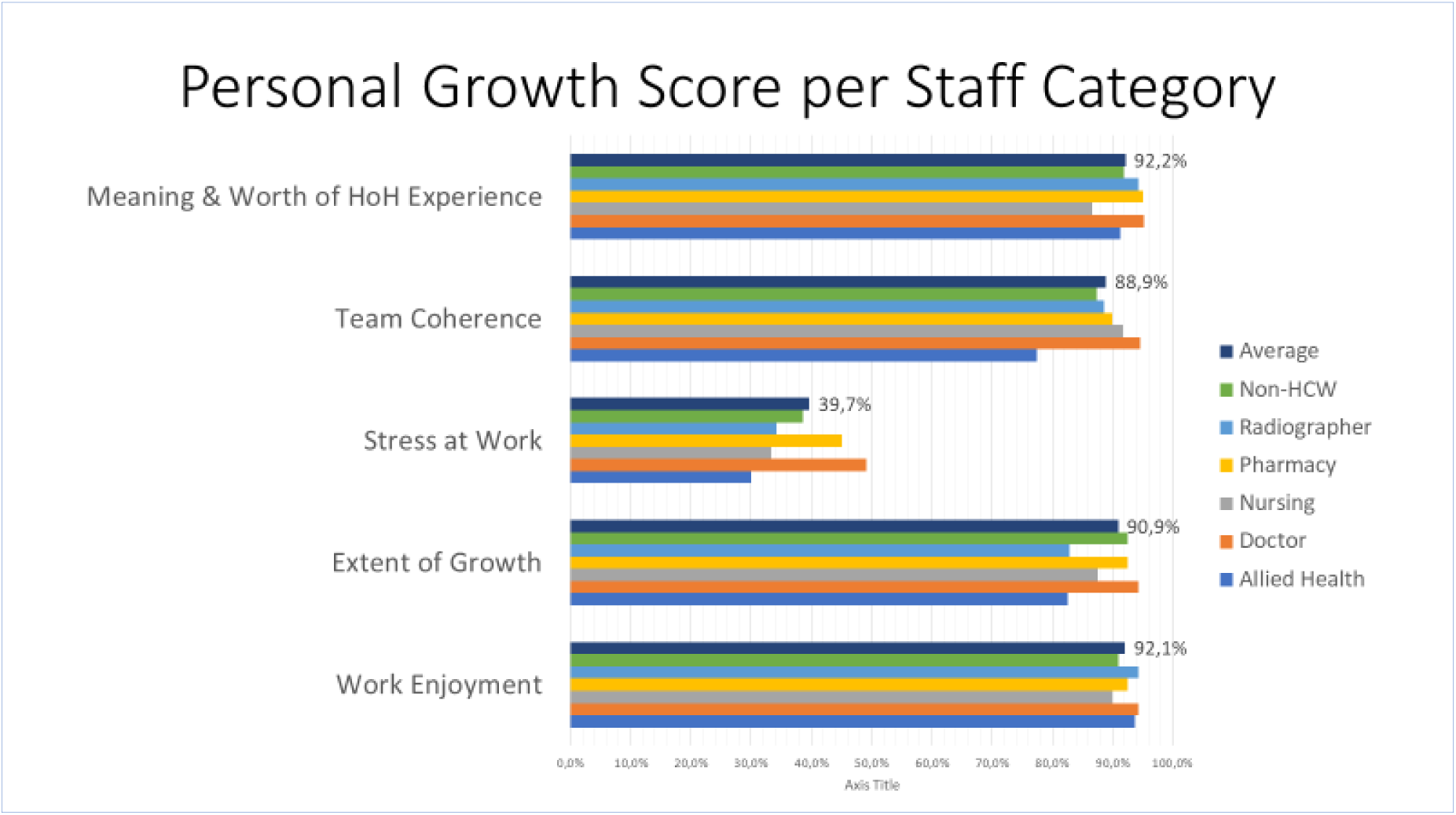
Personal Growth Score per Staff category

In qualitative responses, some expressed their personal growth as follows:

> *“I’ve learned a lot, I know things I’ve never known before, met the most wonderful people and I’ve never known what I want for myself but now I know what I want”*
>
> *“It was so much more than what I was expecting. In learning about the pandemic and growing as a person, finding my strengths”*
>
> *“I did not expect it to be such an amazing and uplifting experience*.*”*

One manager commented on how *“working on the frontline with the constant reminder of COVID… contributed to the development of gratitude and investment in personal relationships with loved ones*.” Others expressed how they learned to be more compassionate, more self-reflective, and how they learned to become better listeners and more effective team players, especially *“when there is such a big hype and when things are moving so rapidly*”. (Manager)

#### d. Meaning

Reflecting on the staff experience, a senior manager highlighted the outcome of hope, which formed an explicit part of the aim as well as the name of the HOH.

> *“There was a sense of hopelessness descending on the system and when the CTICC started accepting patients that sense of hopelessness or that dark cloud was certainly lifted”*.

The expressed intention of the HOH appears to have been vindicated in retrospect, as expressed by staff, the patients and their relatives:

> *“They experienced it as a time of hope and not a time of despair and I think that was remarkable. Looking back and knowing that we could have contributed in that way to someone’s experience in that time of fear and anxiety and uncertainty, I think was remarkable and that is something personally that one can really reflect on*.*”* (Senior manager)

#### e. Lessons for the routine delivery of healthcare

In light of the challenges of the first wave of COVID-19, one respondent provided a novel view of approaching challenges and making mistakes, remarking that, “*Whatever we were doing, we were going to make mistakes… I was going to make mistakes and it was okay, but if I see it as a learning curve it’s going to take away that negative thinking of making a mistake and being hard on myself, but rather trying to see it as it is a learning curve: I must learn from it and then move forward*.*”* (Manager). Despite its short lifespan, Hospital of Hope offered numerous learning opportunities and “aha moments” for individuals and for the system going forward.

The Hospital of Hope demonstrated the importance in moving towards an integrated, collaborative approach in which institutions adapt and respond to the needs of each other within the system, away from the norm in which each healthcare facility functions in isolation largely oblivious to the needs of the other facilities with the system. To use the words of one manager, *“We always say that the Department of Health is not an organization… It is multiple organizations within a massive system and just being able to almost be a bit of a bridge or a glue between some of those various organizations showed me so much that I wouldn’t have learnt in my entire career… [This experience provided] a deeper understanding of how everything is knit together*.” Further, another respondent emphasized that the provision of healthcare *“only works as a system when it’s a collaborative effort like this was and when it’s system-wide where the needs are met across the entire platform*.*”* (Senior clinician).

During the close-out management meeting, the way the daily huddles were conducted at Hospital of Hope was valued. It was cited as a crucial source of information, an effective tool to *“identify where the problems are and where the pressures are within the platform”* and a means by which that information can be communicated promptly and widely. It was hoped that this can be taken forward into routine services as well.

In discussing the way forward and ideas for the routine health system in the future, one manager expressed it most clearly as follows:

> *“This was a very reactive and responsive approach but I mean if we can focus that attention just as we did with this pandemic on something like preventative medicine and health promotion, if some of that effort and focus goes into something like this, then I think we will really see an overhaul of our health system. So the first thing is to be able to clearly identify what are really the health demands and then secondly, being able to respond to that focusing resources, focusing attention, putting people together to really make a difference in those spaces. So to almost get rid of all the clutter. I think the clutter is taking up so much time and really distracting us from what is important and we have really seen now that what is important is where the health system and the communities meet and what is important in the community in the health sector or from a health perspective must also be what is important for us as a health department and we got this right during this pandemic*.*”* (Manager)

## DISCUSSION

The results reveal surprisingly positive outcomes from the perspective of the staff involved in the field hospital, in contrast to the reports from overseas^9,10^ as well as in South Africa^11^, which described stressed and overworked health personnel struggling to keep up with the overwhelming number of patients. Recent research supports these anecdotal reports of increased incidence of burn-out amongst HCW experienced during the COVID-19 pandemic^12-15^.

The CTICC HOH was conceived, planned and implemented in an extraordinarily short space of time of 12 weeks before the first patients arrived on the 8^th^ of June 2020. In contrast to the precedents of the Nightingale Hospitals in the United Kingdom and other temporary facilities established in other countries, the crucial strategic decisions were made surprisingly well in retrospect, considering the degree of uncertainty in modelling at the beginning of the first wave. The managers of the Western Cape Provincial Health department demonstrated extraordinary foresight and leadership in mobilizing the necessary resources and making the right calls on major decisions such as oxygen supply and the number of beds, and a stepwise approach proved wise. For example, the strategic management decision not to extend the facility as planned by a further 400 beds was vindicated by the sudden drop in cases towards the end of July 2020. This could lead to the conclusion that the positive staff outcomes could be regarded as merely a matter of the level of resources being sufficient for the need.

However, other factors contributed also significantly to the positive outcomes, as it has been shown that resources alone are not enough to create a successful health service, particularly in LMICs^16^. Despite an initial disjuncture between the design and the implementation teams, the delegation of the medical management of the facility to family physicians ensured that a person-centred approach pervaded both staff relations and patient care, appropriate to the intermediate level of care, in contrast to the acute level of care where an emergency approach was more appropriate. In an industrial type of setting such as the convention centre with systems established to cope with large numbers of patients, the tendency towards depersonalization was countered by the deliberate focus on relationships, as promoted by the principles of family medicine^17,18^. Thus video-calls with family members at home who were not allowed to visit the patients, were considered as equally important as their medical care.

This focus on relationships extended to staff as well. The motivation of most staff members who were willing to put themselves and their families at risk, in order to play a part in making a difference to the pandemic, carried significant momentum throughout the period of operation of the HOH, and brought staff together around the shared purpose. This enabled an unusual degree of teamwork and cooperation, as attested to by the pharmacists and physiotherapists, for example.

In addition, the responsiveness of the CTICC HoH to the actual needs of the acute care facilities in the Cape Town metropole,^19^ through daily communication across the platform, contributed significantly to the gratitude expressed by the managers, who described the CTICC HoH opening as “*just in time*” to relieve the load.

The study was limited by the response rate to the survey, particularly of nurses, and the relatively few key informant interviews. The exit questionnaire was not piloted before implementation. However, this was mitigated by the multiple sources and types of information collected, including qualitative and quantitative data from the video, documents and interviews.

## CONCLUSION

The staff experience of the first wave at the CTICC Hospital of Hope was unique and unprecedented, and it produced significant learnings for the design and management of routine health services outside of a disaster situation. The staff recruited to the HoH largely volunteered out of a sense of wanting to contribute positively to mitigating the effects of the pandemic, despite the known risks. This has direct implications for recruitment procedures in routine services, suggesting that the fundamental motivations of applicants should be probed. Cultivating a unity of purpose amongst the staff is the primary task of every leader in optimizing the impact of an organization. That this unity of purpose was focused around person-centredness was a deliberate design decision for the intermediate level of care, by appointing family physicians as clinical team leaders. Similarly, the low staff infection rate demonstrated what can be achieved through intelligent design and careful implementation to assure staff of their safety and support. The adaptability and responsiveness of the facility and its staff was largely a product of the unprecedented nature of the pandemic, but such approaches could benefit routine health services enormously, as individual hospitals and health facilities realize their place in a system is ‘more than the sum of its parts’.

## Data Availability

All relevant data are within the manuscript and its Supporting Information files.

## LIST OF ABBREVIATIONS

COVID-19: Coronavirus disease 2019
CTICC: Cape Town International Convention Centre
HoH: ‘Hospital of Hope’

## DECLARATIONS

### Ethics approval and consent to participate

The study was approved by the Human Research Ethics Committee at the University of Cape Town, reference 503/2020.

### Consent for publication

Not applicable.

### Availability of data and materials

The datasets used and/or analysed during the current study are available from the corresponding author on reasonable request.

### Authors’ contributions

SR, KP and TR conceived the study, MN, RO, NH and TA collected the data, SR KP, MN, RO, NH and TA analysed the data and drafted sections of the manuscript. All authors reviewed and amended the manuscript.

## Acknowledgements

All key informant interviewees, and all staff of the CTICC HOH.

## Authors’ information (optional)

SR: BSc(Med), MBChB, MFamMed, PhD – Head of Primary Health Care, University of Cape Town

TA: MPH – Facility Manager, Elsies River Community Health Centre, Western Cape Department of Health

NH: MBChB – Medical Officer, Western Cape Department of Health

MN: BCom (Hons), MCom, CA(SA), MBChB – Medical Officer and Research Assistant, University of Cape Town

RO: MBChB, MMed(FamMed) – Consultant, Western Cape Department of Health

TR: MBChB, MMed(FamMed) – Senior Lecturer, Family Medicine, University of Cape Town

KP: MBChB, MMed(FamMed), FCFP(SA), PhD – Head of Family Medicine, University of Cape Town

All the authors volunteered to work at the CTICC HOH for the duration of the first wave in various positions: TA was the facility manager; TR and KP were clinical managers; SR and RO were consultants; and NH and MN were medical officers.

## APPENDIX 1

Exit survey https://docs.google.com/forms/d/1fzdzwMc0U77iUpaWNXtIBgr6Zu0Nax4qPhgRaXF1Ih8/edit?ts=5fa00ef0&gxids=7757

## APPENDIX 2

Semi-structured interview guide https://docs.google.com/forms/d/1fzdzwMc0U77iUpaWNXtIBgr6Zu0Nax4qPhgRaXF1Ih8/edit?ts=5fa00ef0&gxids=7757

